# Pretransplant and posttransplant erythroferrone levels and outcomes after heart transplantation

**DOI:** 10.64898/2026.02.20.26346755

**Authors:** Barbara Pitta Gros, Angela Rocca, Natallia Laptseva, Michele Martinelli, Andreas Flammer, Henri Lu, Philippe Meyer, Nicolas Leuenberger, Martin Müller, Roger Hullin

## Abstract

**Background:** Iron metabolism disorder is highly prevalent before and after heart transplantation (HTx). The impact of pretransplant and posttransplant iron disorder on posttransplant outcomes is unclear.

**Objective:** Pretransplant serum levels of key regulator proteins of iron metabolism (hepcidin, interleukin-6, erythroferrone) were tested for prediction of the composite outcome 1-year posttransplant all-cause mortality (ACM) or ≥moderate acute cellular rejection (ACR). Furthermore, serum levels of these proteins were measured at 1-year posttransplant to explore their posttransplant course and association with ACR.

**Results:** In a multicenter cohort including 276 consecutive HTx recipients, patients with or without outcome (n=118/158, respectively) did not differ for pretransplant demographics, mismatch of donor/recipient sex, mismatch of HLA epitopes, and hepcidin or interleukin-6 levels. However, pretransplant erythroferrone levels were higher (1.40 vs. 1.19 ng/mL; p=0.013) and hemoglobin levels were lower (124.5 vs. 127 g/L; p=0.004) among patients with the composite outcome. Pretransplant erythroferrone levels >2.25 ng/ml (4^th^-quartile) were significantly associated with the composite outcome in multivariable analysis (OR 2.17; 95% CI 1.19-3.94, p=0.011; reference: 1^st^-3^rd^ quartiles). In adjusted predicted proportions analysis, the incidence of the composite outcome was higher in 4^th^-quartile patients when compared to 1-3^rd^ -quartiles patients (58.0 vs. 37.7%; p=0.003). At 1-year posttransplant, 80.4% of patients with pretransplant erythroferrone levels >2.25 ng/ml remained high; 88.4% of patients with pretransplant erythroferrone levels ≤2.25 ng/ml had high levels posttransplant. In 1-year survivors with high erythroferrone levels and ≥moderate ACR during the first postoperative year, the ratio of the opponent regulators of hepcidin gene expression, erythroferrone to interleukin-6, was higher when compared to those without ACR (1.18 vs. 0.41; p=0.016). Hepcidin levels were not different between these two subgroups indicating disproportionate ERFE increase.

**Conclusion:** High pretransplant erythroferrone levels predict the composite posttransplant outcome 1-year ACM and ACR. Disproportionately high posttransplant erythroferrone levels are related with ≥moderate acute cellular rejection.

## Introduction

HTx still remains the treatment of choice for eligible patients with advanced HF [1]. Iron deficiency is prevalent in up to 80% of patients on the HTx waitlist [2] and half of all HTx recipients in the maintenance phase after transplant surgery [3–5]. This suggests extension of pretransplant iron metabolism disorder into the posttransplant phase, and this hypothesis receives support from the observation that low pretransplant, but not low posttransplant hemoglobin levels are predictive for 1-year ACM [6].

Iron is not only essential for erythropoiesis but also for biosynthesis, DNA synthesis and repair, and host defense [7]. Iron metabolism disorder can therefore underlie a wide variety of pathologies impacting on posttransplant outcomes such as cardiac allograft failure, allograft rejection, or transplant infectious disease [8].

Iron homeostasis was assessed in this study by measure of serum levels of central regulator proteins of iron metabolism, hepcidin, IL-6 and ERFE [9]. Hepcidin is the key regulator protein of the iron metabolism since it blocks export of iron from intracellular stores into the bloodstream by means of inhibitory interaction with the external part of the iron conducting pore, ferroportin, the only iron exporting cellular membrane protein [9]. Gene expression of hepcidin is under control of two distinct intracellular signaling pathways, the Jak/STAT3 pathway and the bone morphogenetic protein (BMP)/Smad pathway, which both increase hepcidin gene expression upon activation. The Jak/STAT3 pathway is activated by IL-6 in settings of inflammation [9] whereas the BMP/Smad pathway is activated by BMP 2 and BMP 6 which signal adequate iron availability in peripheral pools [9]. Activation of BMP/Smad pathway is counterregulated by ERFE [10] which sequesters BMP 2 and BMP 6 from the circulation [11–13] and this even up to an extent that rises hemoglobin levels in patients with anemia of inflammation [9,14]. In addition, ERFE sequesters BMP 7 [11] which maybe of importance after HTx because high BMP 7 levels orient the proinflammatory M1-macrophage towards the anti-inflammatory M2-macrophage [15,16], and, in addition, attenuate interleukin-2 secretion of CD4^+^-T-cells [16]. Overall, this suggests that sequestration of BMP 7 could increase the risk for allograft rejection.

This study investigated the clinical relevance of pretransplant iron homeostasis for the posttransplant outcomes 1-year ACM and ≥moderate ACR. In addition, posttransplant iron homeostasis was explored for its relationship with ≥moderate acute cellular rejection (ACR).

## Materials and Methods

### Study design

This study is a subproject of the multicenter Swiss Transplant Cohort Study (www.stcs.ch, ClinicalTrials.gov Identifier: NCT01204944) which is a national cohort with prospective follow-up of consecutive solid organ transplant recipients in Switzerland.

The STCS cohort study has national ethical approval; the protocol of this subproject (STCS project: FUP 118) was approved by the scientific committee of the STCS and the ethics committee of the canton Vaud (CER VD-2018-02039). All participants gave informed consent prior to the inclusion into the study. The investigation conformed to the principles outlined in the Declaration of Helsinki.

### Setting

The current study included 276 HTx recipients with transplant surgery at 3 HTx centers (University Hospitals of Berne, Lausanne, Zürich) between May 1, 2008, and end of 2017. Immunosuppression was guided by a national protocol which applies routine EMB procurement for rejection surveillance [17] and agrees with the international consensus recommendations for the care of HTx recipients [18].

### Participants

Included were consecutive adult HTx recipients; excluded were HTx recipients with antibody mediated rejection. ACR was graded according to the ISHLT consensus recommendations [19]. At the day of HTx, 114 among the 276 HTx recipients had continuous-flow LVAD treatment, 26 were on ECMO support, and 6 had intraaortic ballon counterpulsation. The total number of EMBs within the 1-year posttransplant was 3397; 2168 EMBs showed no rejection (0R), 1106 EMBs presented mild rejection (1R), and 123 EMBs had moderate or severe ACR (2R, 3R). Immediate posttransplant immunosuppression followed the national protocol [17] and applied at 1-year posttransplant cyclosporine (n=89), tacrolimus (n=117), everolimus (n=51), mycophenolate mofetil (n= 218), methylprednisolone (n=225), and azathioprine (n=10). Within the first posttransplant year, the national protocol proposed trough levels between 200-250 ug/l for cyclosporine, 6-10 ug/l for tacrolimus, and 6-10 ug/l for everolimus. CMV and pneumocystis prophylaxis were applied as described elsewhere [17].

### Variables

The composite outcome comprised ACM or ACR within the first year post-transplant. Incident ACR was considered when EMB presented with grade ≥2R (=moderate or severe ACR) [19] which is of clinical relevance [18] and asks for adjustment of immunosuppression [17,18]. The incidence analysis of the composite outcome was adjusted for identified confounders.

### Data sources

Baseline and 1-year posttransplant parameters were extracted from the STCS data base; subproject related data not documented by the STCS data base were retrieved from individual EHRs by local co-authors (Bern: MM; Lausanne: BPG; Zürich: NL). Blood drawings were executed for the baseline within the 24 hours’ time interval preceding the transplant operation and at 1-year time interval ± 4 weeks to transplant operation. Routine laboratory parameters were measured by local departments of laboratory medicine. ERFE, hepcidin, IL-6, and ferritin levels were determined from frozen serum samples, immediately after thawing. These measures were executed at the Swiss Antidoping Laboratory, Department of Legal Medicine, University Hospital Lausanne, which has ISO/IEC 17025 certification and applies World Anti-Doping Agency standards.

IL-6 and ferritin serum concentrations were quantified by a fully automated immunoassay system (ADVIA Centaur® XP Immunoassay System by Siemens; IL-6 set #016; ferritin set #297; Germany). For serum hepcidin measurement, serum protein was extracted, precipitated, dried, dissolved, and measured by liquid-chromatography high-resolution mass spectrometry (LC-HRMS) as described elsewhere [20]. However, instead of the full scan strategy, single ion monitoring was used to target the hepcidin-25 isoform, the most active isoform, by means of Q Exactive™ Plus Hybrid Quadrupole-Orbitrap™ Mass Spectrometry (ThermoFisher Scientific, Germany) [21]. ERFE serum concentration was measured using the human erythroferrone ELISA kit (AdipoGen® Life Sciences, Erythroferrone (human) ELISA Set (AG-46B-0012S-KI01), Switzerland) [22,23].

### Statistical analysis

Categorical variables were presented as counts (percentages); continuous variables were summarized using median ± interquartile ranges (IQR) or mean ± standard deviation (SD) based on the results of Shapiro-Wilk normality test. For comparison, the Chi-squared test, the Wilcoxon rank-sum test, or the unpaired *t*-test were used as appropriate. Wilcoxon signed-rank test was used for joint sampling analysis. Univariable logistic regression analysis was used to quantify the association between ERFE levels and the composite outcome. For tests investigating the effect of the ratio of ERFE and IL-6, IL-6 levels were always increased by 0.1 to avoid extremely high ratios in patients with low IL-6 levels.

Multivariable analysis of the composite outcome included parameters with univariable association p <0.1 (supplementary table 3) except for IL-6 which was forced into the model based on pathophysiological considerations. The quality of the multivariable logistic regression model was evaluated using the area under the receiver operation characteristic curve (AUROC). Multivariable analysis results were presented as odds ratios (ORs) with 95% confidence intervals (CI).

Using regression analysis, the incidence rate of the composite outcome and its components was estimated in predicted proportions adjusted for confounding variables.

Statistical analyses were conducted using STATA version 18.1 (StataCorp®, College Station, TX, USA), with a p-value <0.05 designated as the threshold for statistical significance.

## Results

### Study population

Median recipient age was 52 years [IQR 25,75%: 41,59 years], the study participants were predominantly male (75.4%).

### Pretransplant demographic, clinical, and biological parameters of HTx recipients without or with the composite outcome

Study participants without or with the composite outcome did not significantly differ with regards to recipient age, donor age, recipient or donor sex, mismatch of donor/recipient sex or HLA-epitopes, and cold ischemia time. Likewise, serum levels of ferritin, IL-6, or hepcidin were not different between groups. Sodium or hemoglobin levels were lower in patients with the composite outcomes (p=0.017, p=0.004; respectively). Study participants with outcomes had higher pretransplant ERFE serum levels (1.40 vs. 1.19 ng/ml, p=0.013) (**Table 1)**. **Supplemental tables 1 and 2** compare pretransplant characteristics between study patients without or with either 1-year ACM or ACR.

**Table 1.** Comparison of pre- and posttransplant parameters in HTx recipients without (n=158) or with primary outcome (n=118)

|  | Total | 1y-ACM / 1y-ACR |  |  |
| --- | --- | --- | --- | --- |
|  | Total (n=276) | No (n=158) | Yes (n=118) | P-value |
| Age [receiver] (y) | 52 (41; 59) | 52 (42; 59) | 52 (39; 59) | 0.770 |
| Female sex [receiver] | 68 (24.6) | 38 (24.1) | 30 (25.4) | 0.793 |
| Age [donor] (y) | 45 (32; 54) | 45.5 (33; 55) | 45 (31; 53) | 0.507 |
| Female sex [donor] | 107 (38.8) | 57 (36.1) | 50 (42.4) | 0.288 |
| Mismatch sex donor/recipient | 83 (30.1) | 41 (25.9) | 42 (35.6) | 0.084 |
| Ferritin pre-HTx (µg/L) | 122 (47; 238) | 117 (45; 232) | 132.5 (49; 247) | 0.564 |
| IL-6 pre-HTx (ng/L) | 2.3 (0.8; 6.6) | 2.1 (0.7; 5.8) | 2.7 (1.0; 8.7) | 0.152 |
| Hepcidine pre-HTx (nM) | 0.5 (0.5; 1.5) | 0.5 (0.5; 1.5) | 0.5 (0.5; 1.2) | 0.382 |
| ERFE pre-HTx (ng/mL) | 1.21 (0.86; 2.25) | 1.10 (0.84; 1.84) | 1.40 (0.91; 2.92) | 0.013 |
| Bilirubin pre-HTx (µmol/L) | 10 (8; 17) | 10 (8; 16) | 10 (7; 17) | 0.987 |
| Creatinine pre-HTx (µmol/L) | 115 (92; 154) | 114 (93; 154) | 118 (91; 155) | 0.804 |
| Sodium pre-HTx (mmol/L) | 138 (136; 140) | 138 (136; 140) | 138 (135; 139) | 0.017 |
| Hemoglobin pre-HTx (g/L) | 126 (109; 138) | 127.5 (114; 141) | 124.5 (102; 134) | 0.004 |
| HLA mismatch |  |  |  | 0.429 |
| 1 | 2 (0.7) | 2 (1.3) | 0 (0.0) |  |
| 2 | 7 (2.5) | 2 (1.3) | 5 (4.2) |  |
| 3 | 24 (8.7) | 12 (7.6) | 12 (10.2) |  |
| 4 | 76 (27.5) | 45 (28.5) | 31 (26.3) |  |
| 5 | 107 (38.8) | 64 (40.5) | 43 (36.4) |  |
| 6 | 60 (21.7) | 33 (20.9) | 27 (22.9) |  |
| Cold ischemia (min) | 159 (128; 192) | 167.5 (130; 195) | 150 (123; 187) | 0.105 |

**Abbreviations:** ACM=all-cause mortality; ACR=acute cellular rejection; ERFE=erythroferrone; HLA=human leukocyte antigen; IL-6=interleukin-6; pre-HTx=pre-heart transplantation; p=p-value.
P-values were obtained using the Wilcoxon rank-sum test for continuous variables (indicated by presentation as median and interquartile range) and the Chi-square test for categorical variables.

**Table 2.**
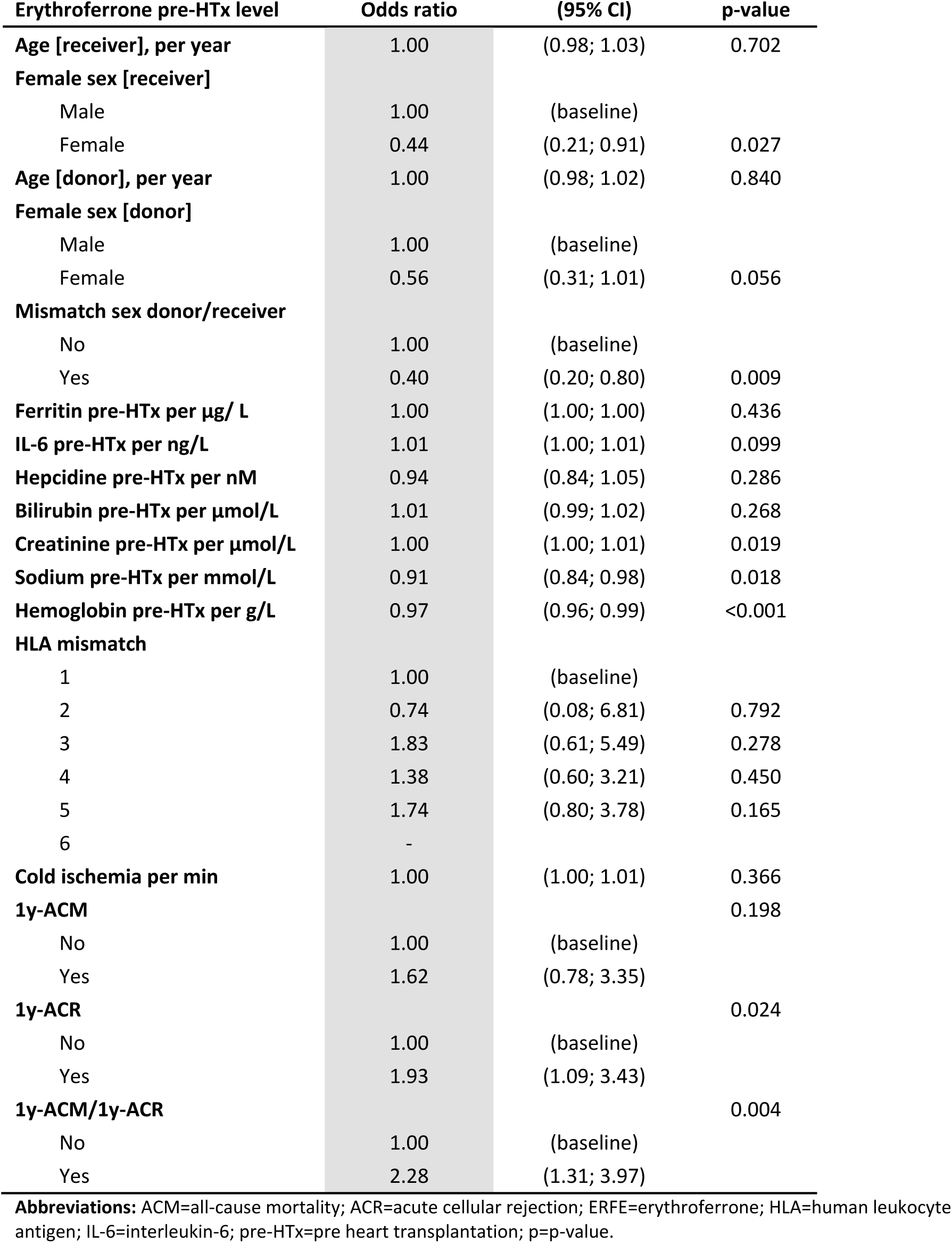
Univariable logistic regression analysis of the association of pretransplant erythroferrone levels and different characteristics.

### Association of pretransplant erythroferrone levels with different parameters

In univariable logistic regression analysis, pretransplant ERFE levels were associated with the composite outcome 1-year ACM or ACR (0.004), 1-year ACR (p=0.024), female sex (p=0.027), mismatch of donor/recipient sex (p=0.009), hemoglobin (p<0.001), sodium (p=0.018), and creatinine (p=0.019) (**Table 2**).

### Comparison of demographic, clinical, and biological parameters between HTx-recipients with pretransplant erythroferrone levels ≤ and >2.25 ng/ml (1^st^-3^rd^ vs. 4^th^ quartiles)

In HTx-recipients with ERFE levels >2.25 ng/ml (4^th^ quartile) when compared to those with ERFE levels ≤2.25 ng/ml (1^st^-3^rd^ quartile), female sex and donor/recipient sex mismatch were more prevalent (p=0.024, p=0.008; respectively); IL-6 levels were higher (p=<0.001), hepcidin levels were lower (p=0.031), ferritin levels were not significantly different, and hemoglobin levels were lower (p<0.001); bilirubin and creatinine levels were significantly higher (p=0.047, p=0.023; respectively) (**Table 3**).

**Table 3.** Comparison of pre- and posttransplant parameters in study participants with pretransplant 1^st^-3^rd^ quartile vs. 4^th^ quartile erythroferrone levels (≤ and > 2.25 ng/mL)

|  | Total | Erythroferrone pre-HTx level |  |  | P-value |
| --- | --- | --- | --- | --- | --- |
| | Total (n=276) | $\leq 2.25$ (n=207) | $> 2.25$ (n=69) | | |
| Age [receiver] (y) | 52 (41; 59) | 52 (40; 59) | 51 (41; 58) |  | 0.868 |
| Female sex [receiver] | 68 (24.6) | 58 (28.0) | 10 (14.5) |  | 0.024 |
| Age [donor] (y) | 45 (32; 54) | 46 (33; 54) | 44 (31; 53) |  | 0.805 |
| Female sex [donor] | 107 (38.8) | 87 (42.0) | 20 (29.0) |  | 0.054 |
| Mismatch sex donor/recipient | 83 (30.1) | 71 (34.3) | 12 (17.4) |  | 0.008 |
| Ferritin pre-HTx (ng/mL) | 122 (47; 238) | 132 (49; 266) | 100 (42; 203) |  | 0.100 |
| IL-6 pre-HTx (ng/L) | 2.3 (0.8; 6.6) | 1.6 (0.6; 4.5) | 6.3 (2.4; 14.3) |  | <0.001 |
| Hepcidine pre-HTx (nM) | 0.5 (0.5; 1.5) | 0.5 (0.5; 1.6) | 0.5 (0.5; 0.8) |  | 0.031 |
| Bilirubin pre-HTx (μmol/L) | 10 (8; 17) | 10 (7; 15) | 11 (9; 18) | | 0.047 |
| Creatinine pre-HTx (μmol/L) | 115 (92; 154) | 113 (89; 144) | 127 (99; 191) | | 0.023 |
| Sodium pre-HTx (mmol/L) | 138 (136; 140) | 138 (136; 140) | 137 (135; 139) |  | 0.066 |
| Hemoglobin pre-HTx (g/L) | 126 (109; 138) | 128 (115; 140) | 114 (101; 127) |  | <0.001 |
| HLA mismatch |  |  |  |  | 0.123 |
| 1 | 2 (0.7) | 0 (0.0) | 2 (2.9) |  |  |
| 2 | 7 (2.5) | 6 (2.9) | 1 (1.4) |  |  |
| 3 | 24 (8.7) | 17 (8.2) | 7 (10.1) |  |  |
| 4 | 76 (27.5) | 58 (28.0) | 18 (26.1) |  |  |
| 5 | 107 (38.8) | 77 (37.2) | 30 (43.5) |  |  |
| 6 | 60 (21.7) | 49 (23.7) | 11 (15.9) |  |  |
| Cold ischemia (min) | 158.5 (127.5; 191.5) | 155 (128; 186) | 166 (127; 200) |  | 0.158 |

**Abbreviations:** ACM=all-cause mortality; ACR=acute cellular rejection; HLA=human leukocyte antigen; IL-6= interleukin-6; pre-HTx=pre heart transplantation. P-values were obtained using the Wilcoxon rank-sum test for continuous variables (indicated by presentation as median and interquartile range) and the Chi-square test for categorical variables.

### Multivariable analysis of associations with the dependent variable composite outcome

Pretransplant parameters presenting association with the composite outcome in univariable analysis (p<0.1) (**supplemental table 3**) entered into the multivariable analysis; IL-6 was not associated in univariate analysis (p=0.7) but was included for pathophysiological consideration. In multivariable analysis, the composite outcome was significantly associated with ERFE levels >2.25 ng/ml (OR 2.17; 95%CI 1.19-3.94, p=0.011), donor/recipient sex mismatch (OR 2.01, 95%CI 1.16-3.49, p=0.013), and hemoglobin per g/l (OR 0.99, 95%CI 0.97-1.00, p=0.017). IL-6 was not significantly related with the composite outcome. The AUROC was 0.677. (**Table 4**).

**Table 4.**
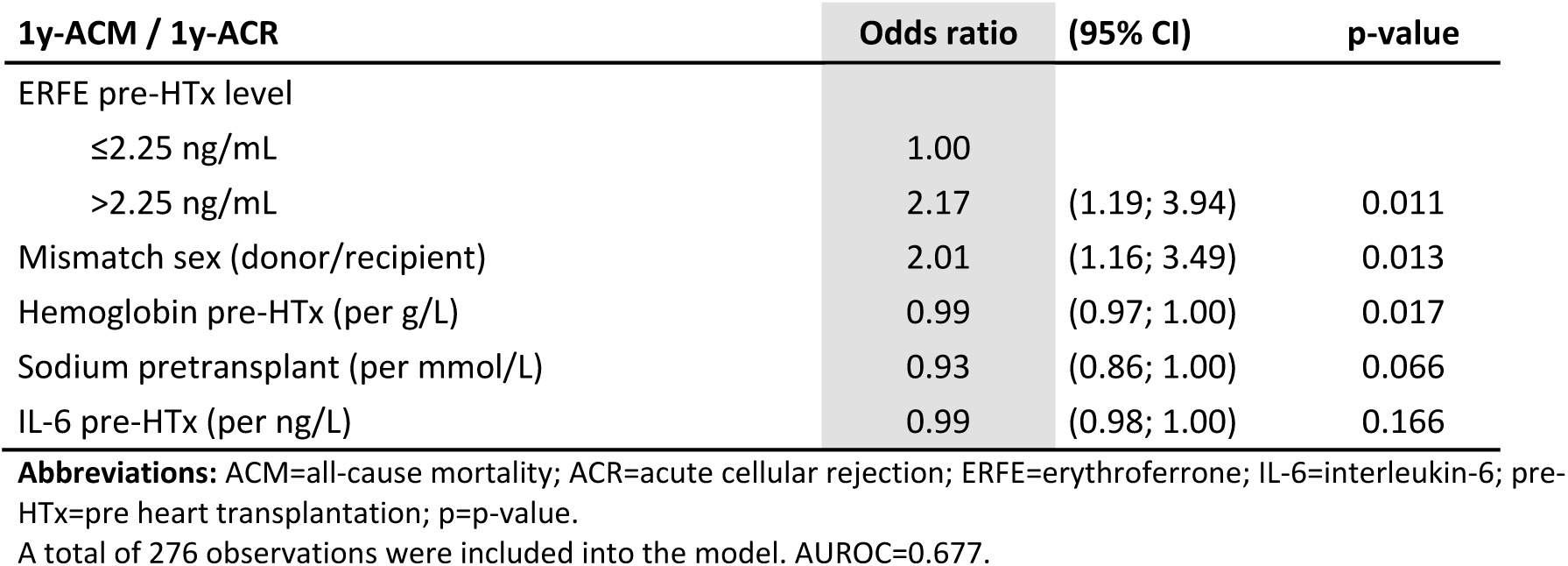
Multivariable logistic regression analysis of parameters associated with the composite endpoint.

| 1y-ACM / 1y-ACR | Odds ratio | (95% CI) | p-value |
| --- | --- | --- | --- |
| ERFE pre-HTx level |  |  |  |
| ≤2.25 ng/mL | 1.00 |  |  |
| >2.25 ng/mL | 2.17 | (1.19; 3.94) | 0.011 |
| Mismatch sex (donor/recipient) | 2.01 | (1.16; 3.49) | 0.013 |
| Hemoglobin pre-HTx (per g/L) | 0.99 | (0.97; 1.00) | 0.017 |
| Sodium pretransplant (per mmol/L) | 0.93 | (0.86; 1.00) | 0.066 |
| IL-6 pre-HTx (per ng/L) | 0.99 | (0.98; 1.00) | 0.166 |

**Abbreviations:** ACM=all-cause mortality; ACR=acute cellular rejection; ERFE=erythroferrone; IL-6=interleukin-6; pre-HTx=pre heart transplantation; p=p-value. A total of 276 observations were included into the model. AUROC=0.677.

### Adjusted predicted proportions for composite outcomes

Patients with pretransplant ERFE levels >2.25 ng/ml had a significantly increased risk for the composite outcome (p=0.011) (**Figure 1**). Adjusted predicted proportions for either 1-year ACM or 1-year ACR were not different between patients with low ERFE levels when compared to patients with high ERFE levels (p=0.16, p=0.093; respectively).

**Figure 1.**
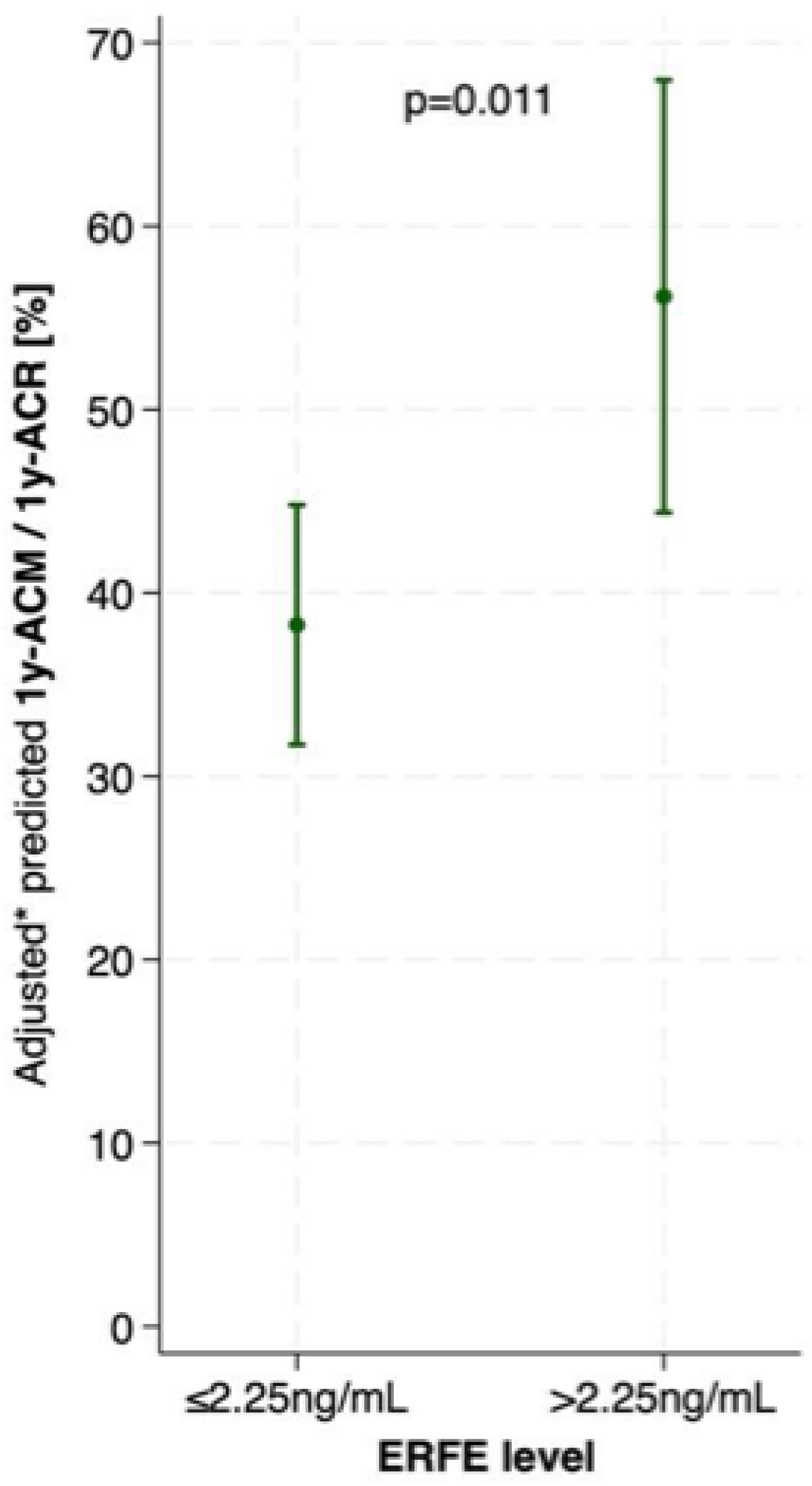
Adjusted predicted proportions for the composite endpoint 1y-ACM and 1y-ACR. *adjusted on hemoglobin, sodium, IL-6 at baseline, and mismatch for sex donor/recipient. Incidence of the composite outcome (n=118). P-value was obtained through multivariable logistic regression analysis.

### Course of ERFE levels between baseline and 1-year posttransplant

88.4% of the 181 1-year survivors with pretransplant 1^st^-3^rd^-quartile ERFE levels increased their 1-year ERFE levels to >2.25 ng/ml. In the 56 survivors with pretransplant ERFE levels >2.25 ng/ml, 80.4% of patients remained with high ERFE levels; 10.7% of patients with high pretransplant ERFE levels decreased their ERFE levels to ≤2.25 ng/ml (**Figure 2**). The Sankey plot in **Supplemental Figure 1** shows patients with 1-year ACM, presents distribution of 1-year survivors into low and high ERFE subgroups, and shows incidence of ACR as a function of low and high ERFE levels at 1-year.

**Figure 2.**
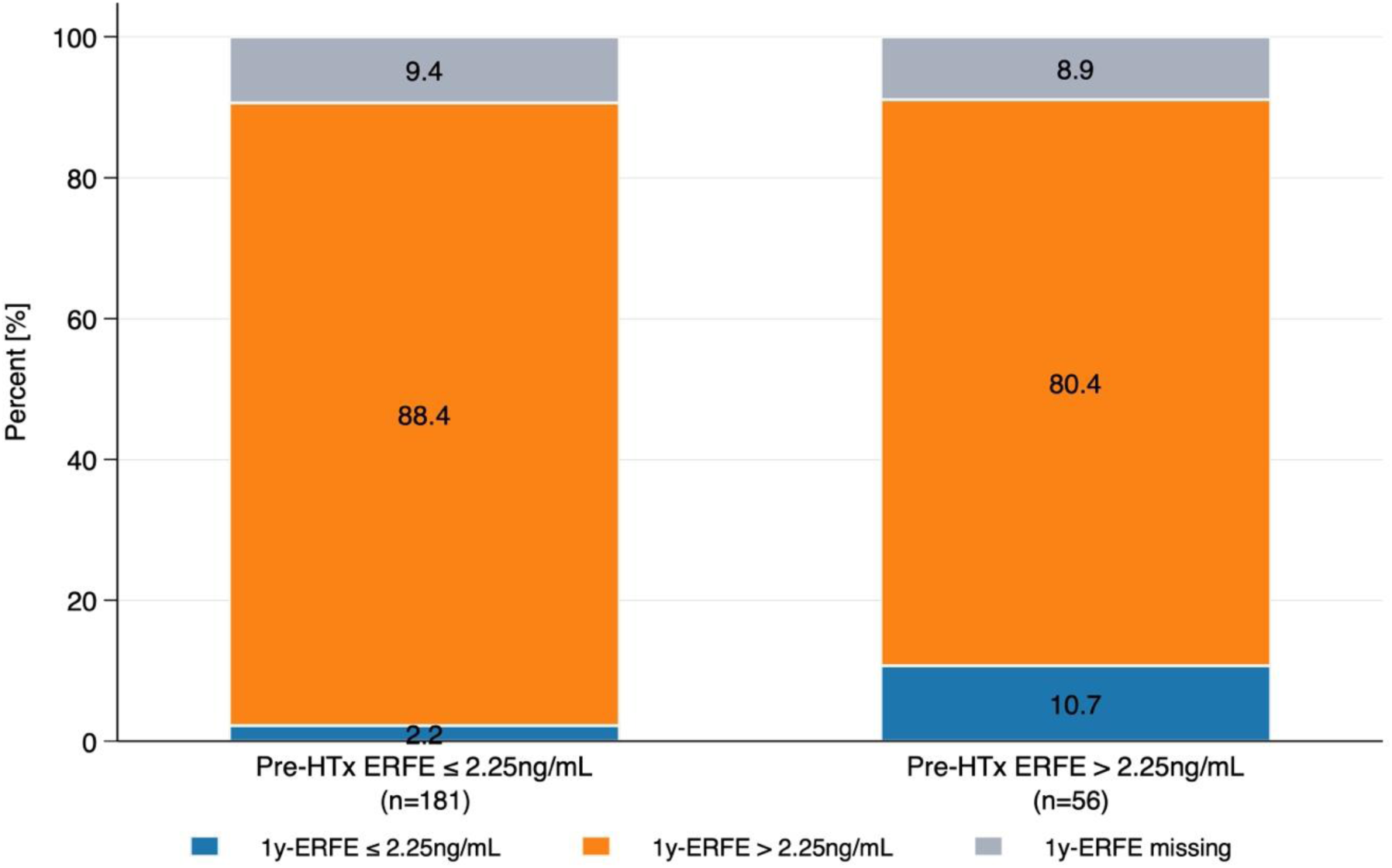
ERFE levels at 1 year after transplantation according to pretransplant ERFE levels. In total, 215 (90.7%) non-missing 1y-ERFE values of 237/276 patients alive 1-year after HTx. **Abbreviations:** ERFE=erythroferrone; pre-HTx=pre-heart transplantation.

### Comparison of the ERFE/IL-6 ratios in subgroups ≤2.25 ng/ml vs. >2.25 ng/ml in patients with or without incident ACR

ERFE/IL-6 ratios were not significantly different between HTx-recipients with pretransplant ERFE levels ≤2.25 ng/ml vs. >2.25 ng/ml when categorized into HTx recipients without or with future ACR (**supplemental figure 2**). Similar results were obtained for the comparison of ERFE/IL-6 ratios in 1-year survivors. However, the ERFE/IL-6 ratio was significantly higher in HTx survivors with high ERFE levels and ACR when compared to HTx recipients with high ERFE levels but without ACR (1.19 vs. 0.41; p=0.016) (**Figure 3**).

**Figure 3.**
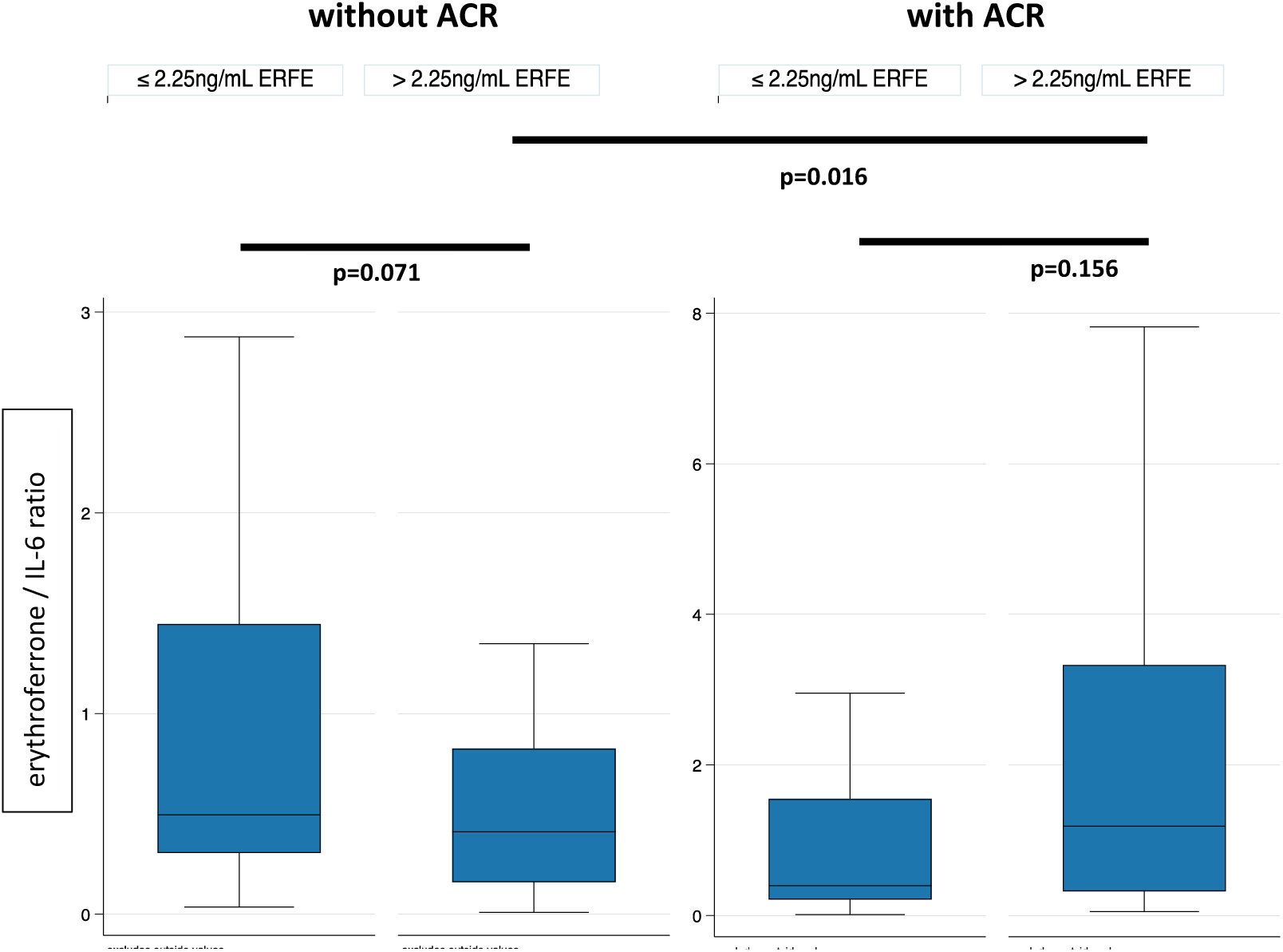
Comparison of the 1-year posttransplant erythroferrone/interleukin-6 ratio in HTx recipients without and with ACR. Patients without ACR: n=115 ≤2.25ng/ml vs. n=28 >2.25ng/ml; patients with ACR: n=49 ≤2.25ng/ml vs. n=23 >2.25ng/ml. P-values were obtained by Wilcoxon rank sum tests.

The following analyses compare regulation of pretransplant iron homeostasis based on hemoglobin, and serum levels of ferritin, hepcidin- and interleukin-6 in patients without or with future ACR at pretransplant (**supplemental figure 2**); posttransplant analyses at 1-year considered without or with incident ACR (**figure 4**). Considering the prediction of the composite outcome in pretransplant patients with ERFE values >2.25 ng/ml, all these investigations were carried out with this cut-off.

**Figure 4.**
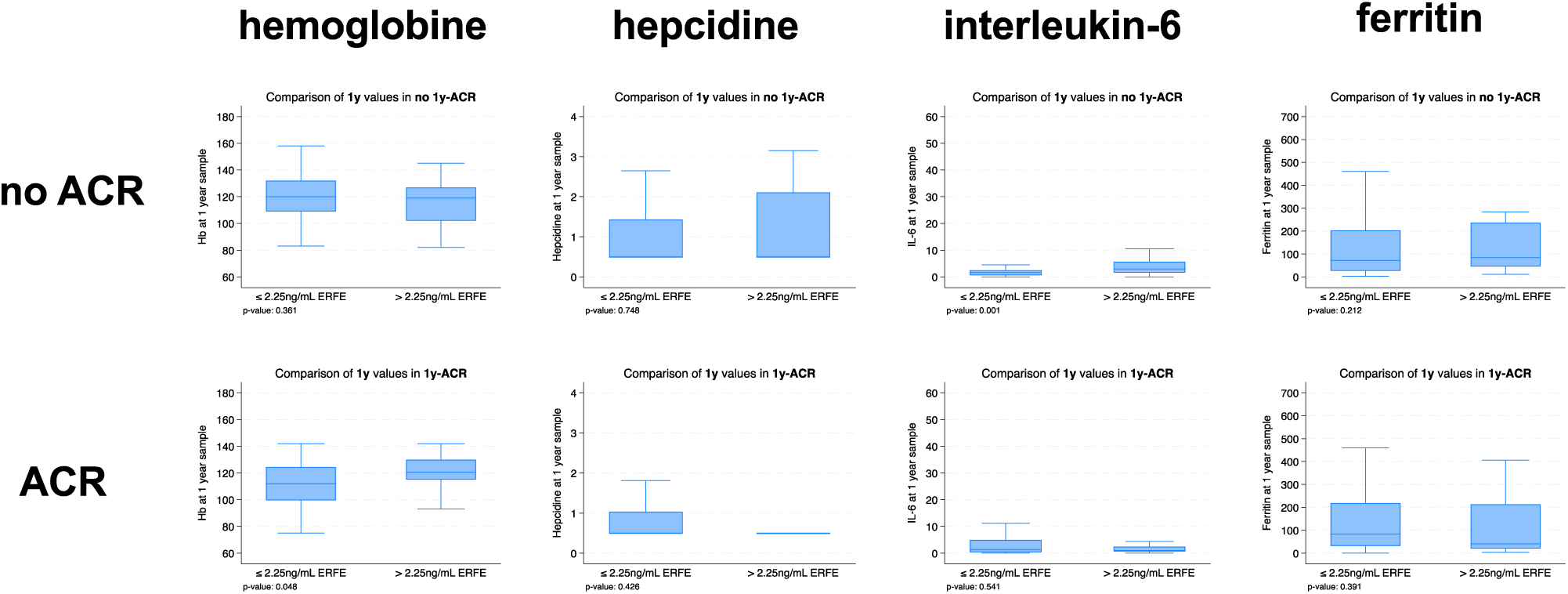
Posttransplant levels of hemoglobin, IL-6, hepcidin and ferritin as a function of ERFE-levels ≤2.25 ng/ml vs. >2.25 ng/ml in patients with or without ACR. ACR=grade 2 or 3 (ISHLT 2004), ERFE=erythroferrone, IL-6=interleukin-6; n=237/276 1y-survivors; no 1-ACR (n=158), ACR (n=79. P-values were obtained with Wilcoxon rank sum tests.

### Pre- and posttransplant levels of hepcidin as a function of ERFE levels ≤2.25 ng/ml vs. >2.25 ng/ml

Pretransplant hepcidin levels were not different when comparing subgroups with low and high ERFE levels in HTx recipients without future ACR (p=0.278) and with future ACR (p=0.050) (**Supplemental Figure 3**). There was no difference at 1-year posttransplant (p=0.748; p=0.426; respectively) (**Figure 4**).

### Pre- and posttransplant levels of interleukin-6 as a function of ERFE levels ≤2.25 ng/ml vs. >2.25 ng/ml

Pretransplant IL-6 levels were low in low-ERFE subgroups and higher in high ERFE-subgroups in HTx recipients without future ACR (1.7 vs. 6.5 ng/l, p<0.001) or with future ACR (1.5 vs. 4.2 ng/l; p=0.002; respectively) (**Supplemental Figure 3**). In HTx survivors without ACR during the first postoperative year, the IL-6 level was lower in the low ERFE subgroup and higher in the high ERFE subgroup (1.6 vs. 3.0 ng/l; p=0.001); in survivors with rejection, IL-6 levels were overall low and not different between patients with low and high ERFE levels (1.0 vs. 1.4 ng/l; p=0.541) (**Figure 4**).

### Pre- and posttransplant levels of hemoglobin as a function of ERFE levels ≤2.25 ng/ml vs. >2.25 ng/ml

In patients without future ACR, pretransplant hemoglobin levels were higher in the low-ERFE subgroup when compared to the high-ERFE level subgroup (131 vs 114 g/l, p<0.001). In patients with future ACR, pretransplant hemoglobin levels were not different between low and high ERFE subgroups (126 vs. 114 g/l, p=0.235) (**Supplemental Figure 3**). At 1-year posttransplant, hemoglobin levels were not different between low and high ERFE subgroups in HTx recipients without ACR (120 vs. 119 g/l, p=0.361); in HTx recipients with ACR during with the first posttransplant year hemoglobin levels were higher in the high ERFE subgroup when compared to the low ERFE subgroup (121 vs. 112 g/l; p=0.048) (**Figure 4**).

### Pre- and posttransplant levels of ferritin as a function of ERFE levels ≤2.25 ng/ml vs. >2.25 ng/ml

Pretransplant ferritin levels were not different between ERFE subgroups in HTx recipients without future ACR (127 vs. 115 ug/l, p=0.580) and lower in the high ERFE subgroup in HTx recipients with future ACR (163 vs. 75 ug/l, p=0.035) (**Supplemental Figure 3**). At 1-year posttransplant, ferritin levels were not different between ERFE subgroups regardless of the absence or incidence of ACR (72 vs. 84 ug/l, p=0.212; 83 vs. 41 ug/l, p=0.391) (**Figure 4**).

## Discussion

High pretransplant ERFE levels were predictive for the composite outcome posttransplant 1-year ACM or ≥moderate ACR in this multicenter observational study cohort. At 1-year posttransplant, the percentage of HTx recipients with high ERFE levels was overall substantially higher when compared to pretransplant. This increase was due to rise of ERFE levels in many HTx recipients with low pretransplant ERFE levels. In HTx survivors with high ERFE levels and incident ACR, the ERFE/IL-6 ratio, which illustrates antagonistic regulation of hepcidin gene expression by ERFE and IL-6, was significantly higher when compared to those without incident ACR. Hepcidin and hemoglobin levels were not different between these groups suggesting relationship between disproportionate ERFE increase and ACR.

Iron status is routinely assessed in the bloodstream of chronic stable HF patients by measure of ferritin and iron saturation of transferrin, in accordance with the HF guidelines [24,25]. However, serum levels of the acute phase proteins ferritin and transferrin vary considerably in advanced HF patients [26,27] and in settings of systemic inflammation such as early after HTx. In addition, corticosteroid treatment, which is routinely applied in HTx recipients in the early posttransplant phase, changes gene expression of these acute phase proteins [28]. Therefore, this study assessed the state of iron metabolism by measure of serum levels of hepcidin, IL-6, ERFE, which allowed for unbiased comparison between pre- and posttransplant measures.

Low pretransplant hemoglobin levels had been predictive for 1-year ACM in an observational monocenter cohort study following HTx recipients [6], and, in concordance, pretransplant hemoglobin levels were also predictive for the composite outcome of the current study. This converging evidence suggests that the key finding of the present study, the prediction of the composite posttransplant outcome by high pretransplant ERFE levels, should be broadly valid.

Before transplant operation, one quarter of all study participants presented high ERFE levels, however, at 1-year after transplant operation, the percentage of HTx recipients with high ERFE levels was substantially higher. This increase was due to the rise of ERFE levels in 88.4% of HTx survivors with low pretransplant levels, in addition, 80.4% of study participants with high pretransplant ERFE levels remained with high ERFE levels. Among these patients, the ERFE/IL-6 ratio was higher in HTx survivors with incident ACR when compared to HTx survivors with high ERFE without ACR. Since hemoglobin and hepcidin levels were not different between these two groups, the increase of ERFE levels in HTx survivors with incident ACR is disproportionate with regard to requirements of the erythron. And this disproportionality applies likewise to IL-6, which showed a low serum level that was not different to the IL-6 values in the subgroup of HTx survivors with incident ACR but low ERFE values. In contrast, the correlation of the disproportionately elevated ERFE and ACR suggests interaction with the adaptive immune system very likely on the bases of the high-affinity sequestration of BMP 2 and BMP 7 [11]. BMP 2 and BMP 7 orient the pro-inflammatory monocyte, or M1 macrophage, towards the anti-inflammatory M2 macrophage [15,16]. In addition, BMP 7 attenuates IL-2 secretion of the CD4^+^ T-lymphocyte which is the immune cell orchestrating allograft rejection and IL-2 [16,29,30]. Excessive sequestration of BMP 2 and BMP 7 could therefore explain the observation that a disproportionate ERFE/IL-6 ratio is associated with ≥moderate ACR.

However, the cause for the large increase of the number of patients with high ERFE levels at 1-year posttransplant remains not entirely clear especially since the number of patients with high ERFE levels was in the current study greater than the number of patients with incident moderate or severe ACR. Both clinical and animal studies provide evidence of a pathophysiological role of high ERFE in settings of cardiovascular disease associated with cardiac injury. In patients with acute dyspnea, high ERFE levels discriminate cardiac origin from other cause [31]. Furthermore, ERFE levels were significantly increased in chronic HF patients with high risk of cardiovascular death and HF hospitalization as shown in a prespecified post-hoc analysis of the EMPEROR reduced and the EMPEROR preserved trials [32]. In addition, high ERFE levels mitigated cardiac injury in animal models of ischemia-reperfusion injury or transaortic constriction [33,34]. Altogether, these observations suggest a cardioprotective effect of high ERFE levels in settings of cardiac injury and this may also explain why patients with type 2 diabetes [35] or dyslipidemia [36] present high ERFE levels. After HTx, HLA mismatch between the donor and the recipient induces immunogenicity-related cardiac injury which maybe an important cause for the large increase of the number of study participants with high posttransplant ERFE levels. On the other hand, the extent of cardiac injury should not be similar for all cardiac allografts because immunogenicity of the cardiac allografts varies as a function of number of mismatching HLA molecules, their capacity to trigger an immune response, and mismatch of donor/recipient sex. The current study provides indirect evidence for this hypothesis that cardiac injury is a cause for high ERFE levels since high pretransplant ERFE levels decreased in 10.7% of HTx survivors to low posttransplant ERFE levels and low pretransplant ERFE levels remained low in 11.6% of HTx survivors.

## Limitations

This study has limitations due to its observational design and the fact that measure of serum parameters of iron metabolism was limited to one pretransplant and another serum sample at 1-year posttransplant. More measures between these two time points would have been helpful to understand better the trajectory of ERFE levels within the first posttransplant year, in particular in HTx recipients with case fatality.

## Conclusion

The clinical significance of ERFE has been seen almost exclusively in connection with erythropoiesis until now. However, results of this and other studies suggest that ERFE not only has the potential to serve as a biomarker for HF severity, but also to become a therapeutic target.

## Data Availability

The data that support the findings of this study are available in the manuscript and in the supplementary material. Additional information is available on reasonable request.

## Further information

## Acknowledgments

We thank the local coordinator of the STCS, Emanuelle Catana, for her support during the data extraction from the respective data bank.

## Data Availability Statement

The data that support the findings of this study are available in the manuscript and in the supplementary material. Additional information is available.

## Abbreviations

ACM: all-cause mortality
ACR: acute cellular rejection
BMP: bone morphogenetic protein
EHR: electronic health record
EMB: endomyocardial biopsy
ERFE: erythroferrone
HF: heart failure
HTx: orthotopic heart transplantation
IL-6: interleukin-6
STCS: Swiss Transplant Cohort Study

---

**Supplemental Table 1.** Comparison of pretransplant parameters of HTx recipients without (n=237) or with 1y-ACM (n=39).

|  | Total | 1y-ACM |  |  |
| --- | --- | --- | --- | --- |
|  | Total (n=276) | No (n=237) | Yes (n=39) | P-value |
| Age [receiver] (y) | 52 (41; 59) | 51 (41; 58) | 57 (41; 62) | 0.023 |
| Female sex [receiver] | 68 (24.6) | 61 (25.7) | 7 (17.9) | 0.295 |
| Age [donor] (y) | 45 (32; 54) | 45 (31; 53) | 48 (43; 55) | 0.025 |
| Female sex [donor] | 107 (38.8) | 87 (36.7) | 20 (51.3) | 0.083 |
| Mismatch sex don/rec | 83 (30.1) | 66 (27.8) | 17 (43.6) | 0.047 |
| Ferritin pre-HTx (µg/L) | 122 (47.5; 238) | 115 (45; 236) | 146 (56; 243) | 0.296 |
| IL-6 pre-HTx (ng/L) | 2.3 (0.8; 6.6) | 2.2 (0.7; 6.3) | 4.1 (1.2; 14.3) | 0.036 |
| Hepcidine pre-HTx (nM) | 0.5 (0.5; 1.5) | 0.5 (0.5; 1.5) | 0.5 (0.5; 1.2) | 0.894 |
| Bilirubin pre-HTx (µmol/L) | 10 (8; 17) | 10 (8; 16) | 11 (9; 21) | 0.149 |
| Creatinine pre-HTx (µmol/L) | 115 (92; 154) | 113 (91; 154) | 126 (95; 161) | 0.178 |
| Sodium pre-HTx (mmol/L) | 138 (136; 140) | 138 (136; 140) | 138 (135; 140) | 0.403 |
| Hemoglobin pre-HTx (g/L) | 126 (109; 138) | 126 (110; 137) | 117 (104; 141) | 0.482 |
| HLA mismatch |  |  |  |  |
| 1 | 2 (0.7) | 2 (0.8) | 0 (0.0) |  |
| 2 | 7 (2.5) | 6 (2.5) | 1 (2.6) |  |
| 3 | 24 (8.7) | 20 (8.4) | 4 (10.3) |  |
| 4 | 76 (27.5) | 62 (26.2) | 14 (35.9) |  |
| 5 | 107 (38.8) | 95 (40.1) | 12 (30.8) |  |
| 6 | 60 (21.7) | 52 (21.9) | 8 (20.5) | 0.793 |
| Cold ischemia (min) | 159 (128; 192) | 158 (128; 192) | 170 (123; 191) | 0.813 |
| Erythroferrone pre-HTx (ng/mL) | 1.21 (0.86; 2.25) | 1.14 (0.85; 2.18) | 1.57 (0.91; 3.19) | 0.106 |

**Abbreviations:** ACM=all-cause mortality; ACR=acute cellular rejection; HLA=human leukocyte antigen; IL-6=interleukin-6; pre-HTx=pre-heart transplantation; p=p-value; y=year. P-values were obtained using the Wilcoxon rank-sum test for continuous variables (indicated by presentation as median and interquartile range) and the Chi-square test for categorical variables.

**Supplemental Table 2.** Comparison of pretransplant parameters of HTx recipients without (n=194) or with 1y-ACR (n=82).

|  | Total | 1y-ACR |  |  |
| --- | --- | --- | --- | --- |
|  | Total (n=276) | No (n=194) | Yes (n=82) | P-value |
| Age [receiver] (y) | 52 (41; 59) | 53 (42; 60) | 50.5 (38; 56) | 0.094 |
| Female sex [receiver] | 68 (24.6) | 45 (23.2) | 23 (28.0) | 0.393 |
| Age [donor] (y) | 45 (32; 54) | 47 (34; 55) | 44 (25; 51) | 0.027 |
| Female sex [donor] | 107 (38.8) | 76 (39.2) | 31 (37.8) | 0.831 |
| Mismatch sex don/rec | 83 (30.1) | 57 (29.4) | 26 (31.7) | 0.700 |
| Ferritin pre-HTx (µg/L) | 122 (47; 238) | 123 (47; 236) | 107.5 (48; 247) | 0.945 |
| IL-6 pre-HTx (ng/L) | 2.3 (0.8; 6.6) | 2.2 (0.8; 6.5) | 2.3 (0.7; 6.7) | 0.992 |
| Hepcidine pre-HTx (nM) | 0.5 (0.5; 1.5) | 0.5 (0.5; 1.5) | 0.5 (0.5; 1.3) | 0.291 |
| Bilirubin pre-HTx (µmol/L) | 10 (8; 17) | 10 (9; 17) | 10 (6; 15) | 0.174 |
| Creatinine pre-HTx (µmol/L) | 115 (92; 154) | 116 (94; 154) | 114 (91; 155) | 0.657 |
| Sodium pre-HTx (mmol/L) | 138 (136; 140) | 138 (136; 140) | 137 (135; 139) | 0.039 |
| Hemoglobin pre-HTx (g/L) | 126 (109; 138) | 127 (112; 141) | 125 (100; 132) | 0.005 |
| HLA mismatch |  |  |  | 0.339 |
| 1 | 2 (0.7) | 2 (1.0) | 0 (0.0) |  |
| 2 | 7 (2.5) | 3 (1.5) | 4 (4.9) |  |
| 3 | 24 (8.7) | 16 (8.2) | 8 (9.8) |  |
| 4 | 76 (27.5) | 58 (29.9) | 18 (22.0) |  |
| 5 | 107 (38.8) | 76 (39.2) | 31 (37.8) |  |
| 6 | 60 (21.7) | 39 (20.1) | 21 (25.6) |  |
| Cold ischemia (min) | 158.5 (127.5; 191.5) | 167 (128; 195) | 150 (127; 185) | 0.072 |
| Erythroferrone pre-HTx (ng/mL) | 1.21 (0.86; 2.25) | 1.16 (0.85; 2.03) | 1.26 (0.88; 2.80) | 0.151 |

**Abbreviations:** ACR=acute cellular rejection; HLA=human leukocyte antigen; IL-6=interleukin-6; pre-HTx=pre-heart transplantation; p=p-value; y=year.
P-values were obtained using the Wilcoxon rank-sum test for continuous variables (indicated by presentation as median and interquartile range) and the Chi-square test for categorical variables.

**Supplemental table 3.** Univariable logistic regression analysis for pretransplant parameters with the composite outcome (n=276).

| 1y-ACM / 1y-ACR | OR | (95% CI) | p-value |
| --- | --- | --- | --- |
| **Age [receiver], per year** | 1.00 | (0.98; 1.01) | 0.693 |
| **Female sex [receiver]** |  |  |  |
| Male | 1.00 | (base) |  |
| Female | 1.08 | (0.62; 1.87) | 0.793 |
| **Age [donor], per year** | 0.99 | (0.98; 1.01) | 0.496 |
| **Female sex [donor]** |  |  |  |
| Male | 1.00 | (base) |  |
| Female | 1.30 | (0.80; 2.12) | 0.289 |
| **Mismatch sex donor/receiver** |  |  |  |
| No | 1.00 | (base) |  |
| Yes | 1.58 | (0.94; 2.65) | 0.085 |
| **Ferritin pre-HTx, per µg/L** | 1.00 | (1.00; 1.00) | 0.673 |
| **IL-6 pre-HTx, per ng/L** | 1.00 | (0.99; 1.01) | 0.700 |
| **Hepcidine pre-HTx, per nM** | 1.00 | (0.95; 1.06) | 0.921 |
| **Bilirubin pre-HTx, per µmol/L** | 1.01 | (0.99; 1.03) | 0.188 |
| **Creatinine pre-HTx, per µmol/L** | 1.00 | (1.00; 1.00) | 0.503 |
| **Sodium pre-HTx, per mmol/L** | 0.92 | (0.85; 0.98) | 0.017 |
| **Hemoglobin pre-HTx, per g/L** | 0.98 | (0.97; 0.99) | 0.004 |
| **HLA mismatch** |  |  |  |
| 1 | 1.00 | (base) |  |
| 2 | 3.06 | (0.55; 17.0) | 0.202 |
| 3 | 1.22 | (0.47; 3.15) | 0.678 |
| 4 | 0.84 | (0.42; 1.67) | 0.622 |
| 5 | 0.82 | (0.43; 1.56) | 0.546 |
| **Cold ischemia time, per min** | 1.00 | (0.99; 1.00) | 0.155 |
| **ERFE pre-HTx, per ng/L** | 1.20 | (1.02; 1.40) | 0.023 |
| **ERFE pre-HTx** |  |  |  |
| ≤ 2.25 ng/mL | 1.00 | (base) |  |
| > 2.25 ng/mL | 2.28 | (1.31; 3.97) | 0.004 |

**Abbreviations:** ACM=all-cause mortality; ACR=acute cellular rejection; ERFE=erythroferrone; HLA=human leukocyte antigen; IL-6=interleukin-6; OR=odds ratio; pre-HTx=pre-heart transplantation; p=p-value.

**Supplemental Figure 1.**
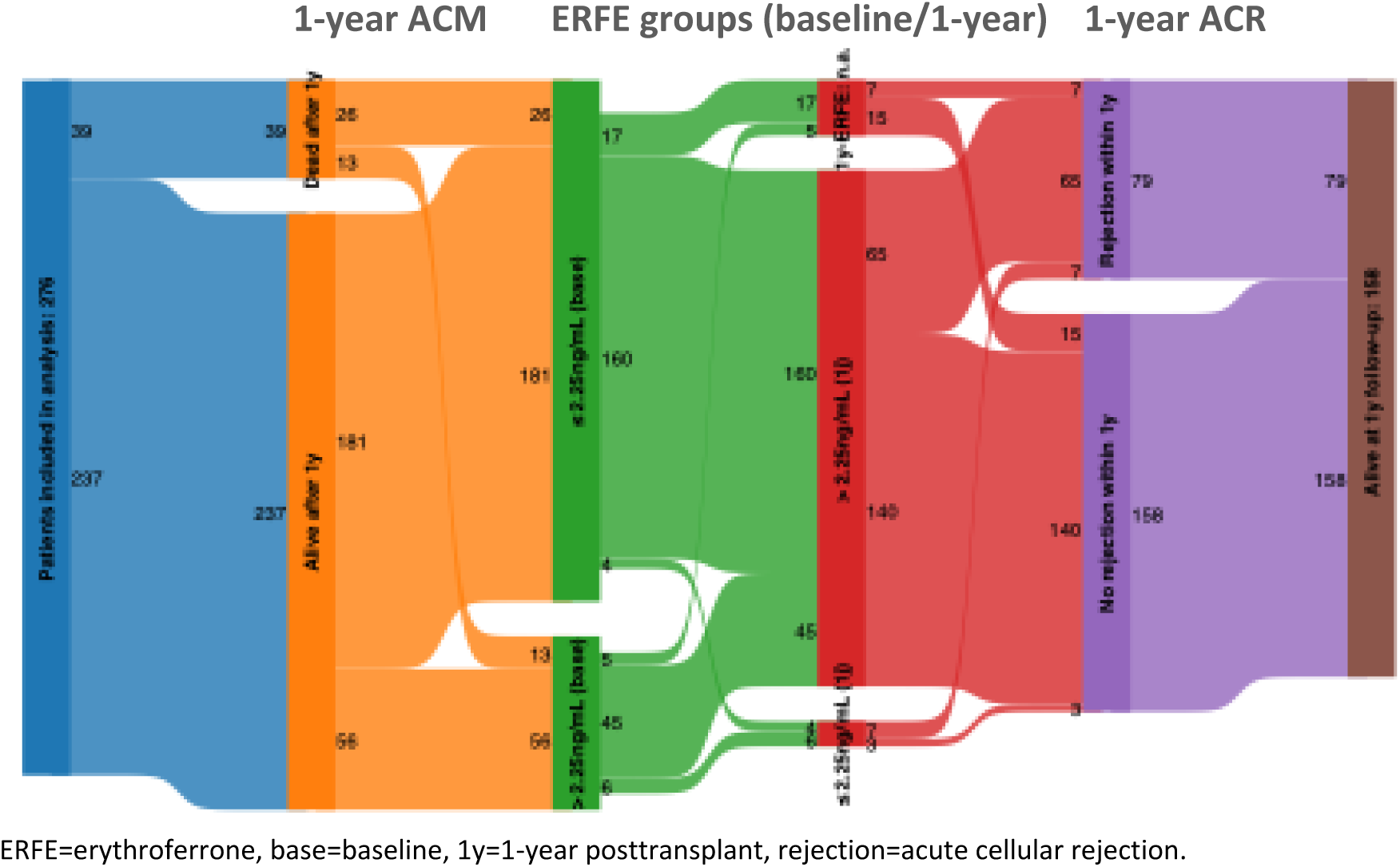
Sankey plot showing the course of 276 study participants as a function of 1-year ACM, ERFE subgroups, and 1-year incidence of ACR.

**Supplemental figure 2.**
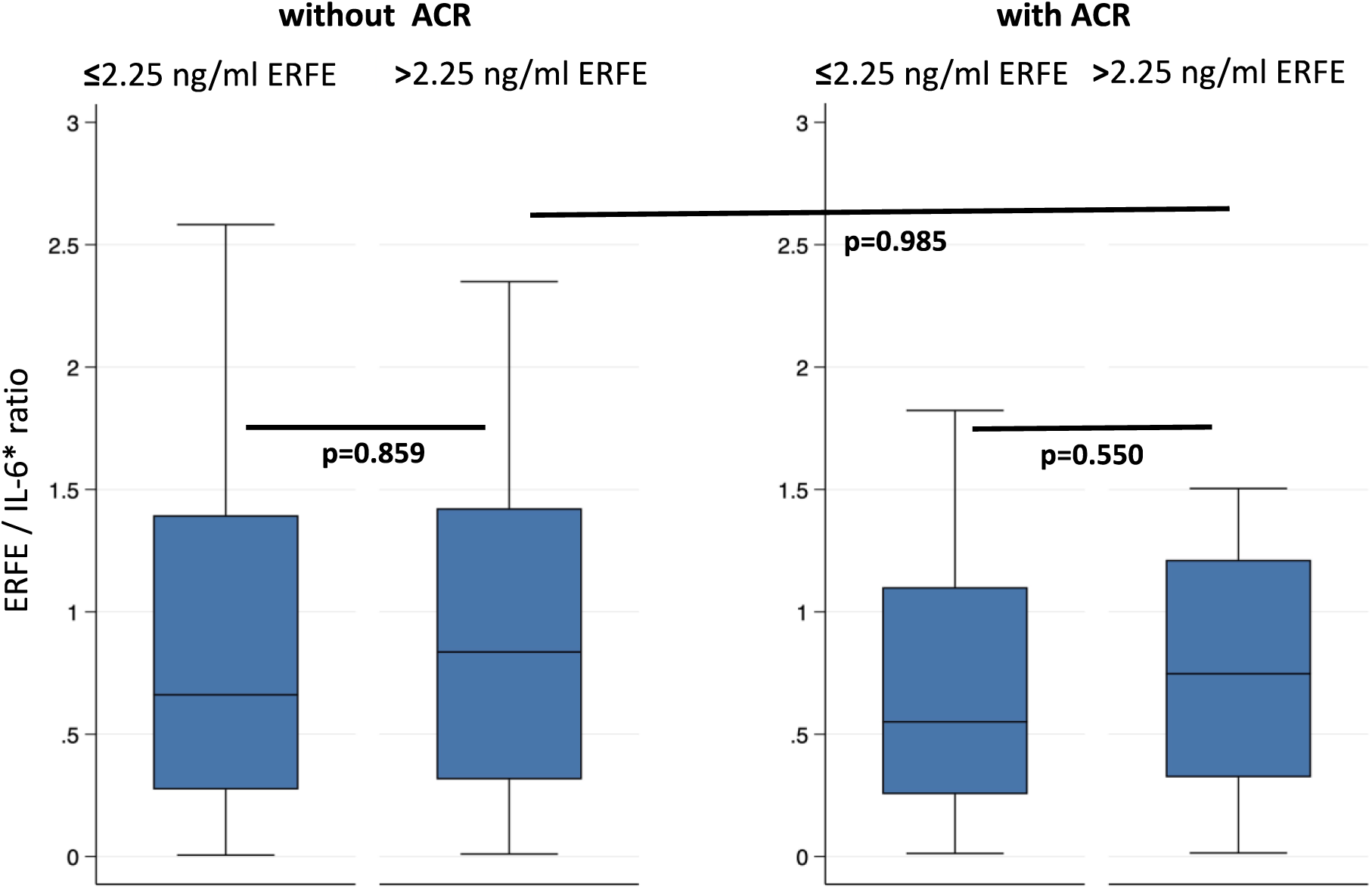
Comparison of the pretransplant ERFE/interleukin-6 ratio in 1y-HTx survivors without and with incident ACR. Patients without missing 1-year posttransplant erythroferrone/interleukin-6 ratio and without ACR: n=143 (≤2.25ng/mL: n= 115; >2.25ng/mL: n=28), with ACR: n=72 (≤2.25ng/mL: n= 49; >2.25ng/mL: n=23). P-values were obtained by Wilcoxon rank sum tests.

**Supplemental Figure 3.**
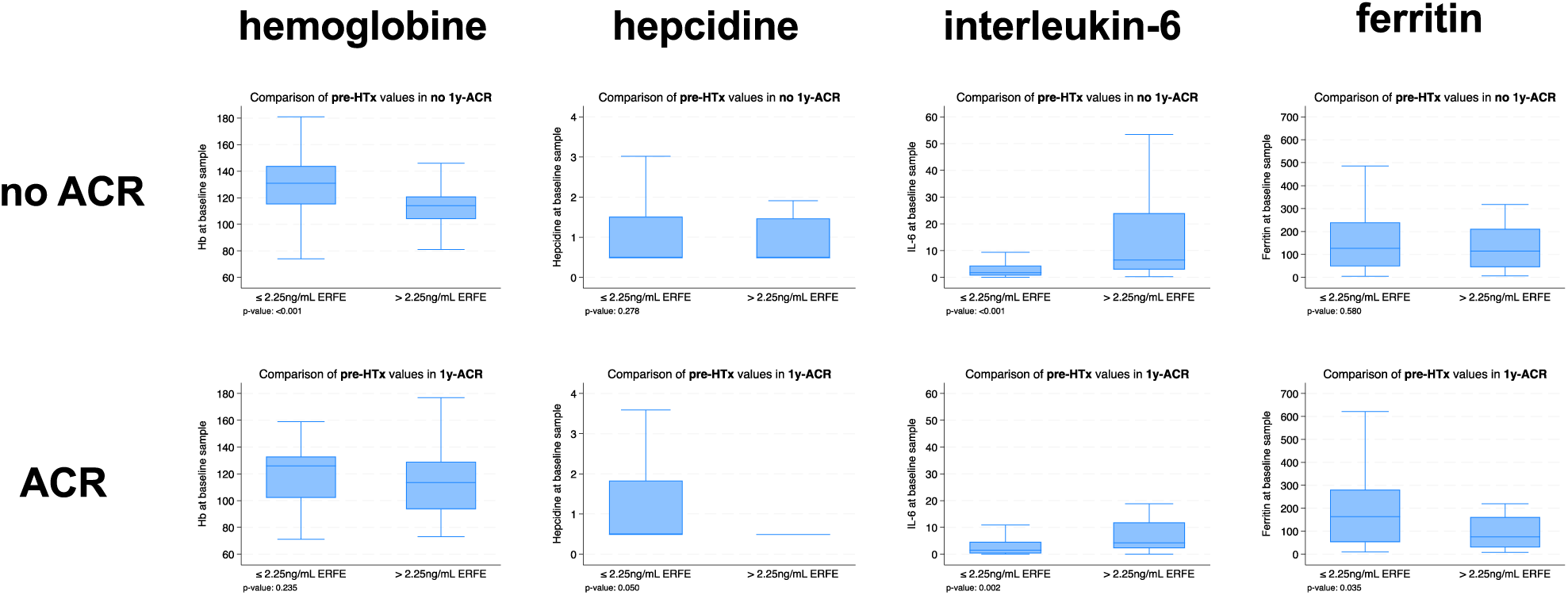
Pretransplant levels of hemoglobin, IL-6, hepcidin and ferritin as a function of ERFE-levels 2.25 ng/ml vs. >2.25 ng/ml in patients with or without ACR (n=276) ACR=grade 2 acute cellular rejection (ISHLT 2004), ERFE=erythroferrone, IL-6=interleukin-6; no ACR (n=194), ACR (n=82). P-values were obtained with Wilcoxon rank sum tests.

